# Locus coeruleus integrity and the effect of atomoxetine on response inhibition in Parkinson’s disease

**DOI:** 10.1101/2020.09.03.20176800

**Authors:** Claire O’Callaghan, Frank H. Hezemans, Rong Ye, Catarina Rua, P. Simon Jones, Alexander G. Murley, Negin Holland, Ralf Regenthal, Kamen A. Tsvetanov, Noham Wolpe, Roger A. Barker, Caroline H. Williams-Gray, Trevor W. Robbins, Luca Passamonti, James B. Rowe

## Abstract

Cognitive decline is a common feature of Parkinson’s disease, and many of these cognitive deficits fail to respond to dopaminergic therapy. Therefore, targeting other neuromodulatory systems represents an important therapeutic strategy. Among these, the locus coeruleus-noradrenaline system has been extensively implicated in response inhibition deficits. Restoring noradrenaline levels using the noradrenergic reuptake inhibitor atomoxetine can improve response inhibition in some patients with Parkinson’s disease, but there is considerable heterogeneity in treatment response. Accurately predicting the patients who would benefit from therapies targeting this neurotransmitter system remains a critical goal, in order to design the necessary clinical trials with stratified patient selection to establish the therapeutic potential of atomoxetine. Here, we test the hypothesis that integrity of the noradrenergic locus coeruleus explains the variation in improvement of response inhibition following atomoxetine. In a double-blind placebo-controlled randomised crossover design, 19 people with Parkinson’s disease completed an acute psychopharmacological challenge with 40 mg of oral atomoxetine or placebo. A stop-signal task was used to measure response inhibition, with stop-signal reaction times obtained through hierarchical Bayesian estimation of an ex-Gaussian race model. Twenty-six control subjects completed the same task without undergoing the drug manipulation. In a separate session, patients and controls underwent ultra-high field 7T imaging of the locus coeruleus using a neuromelanin-sensitive magnetisation transfer sequence. The principal result was that atomoxetine improved stop-signal reaction times in those patients with lower locus coeruleus integrity. This was in the context of a general impairment in response inhibition, as patients on placebo had longer stop-signal reaction times compared to controls. We also found that the caudal portion of the locus coeruleus showed the largest neuromelanin signal decrease in the patients compared to controls. Our results highlight a link between the integrity of the noradrenergic locus coeruleus and response inhibition in Parkinson’s disease patients. Furthermore, they demonstrate the importance of baseline noradrenergic state in determining the response to atomoxetine. We suggest that locus coeruleus neuromelanin imaging offers a marker of noradrenergic capacity that could be used to stratify patients in trials of noradrenergic therapy and to ultimately inform personalised treatment approaches.

## Introduction

Cognitive decline in Parkinson’s disease remains an ongoing therapeutic challenge. The mainstay dopaminergic therapies often fail to improve cognitive deficits, and in some cases can exacerbate them (Cools *et al*., 2003; Lewis *et al*., 2005; Kehagia *et al*., 2010). This has prompted a shift of focus towards other neuromodulatory systems that are affected by Parkinson’s disease and related to cognitive decline, including noradrenaline (Halliday *et al*., 2014). The noradrenergic locus coeruleus is one of the earliest sites of alpha-synuclein pathology (Braak *et al*., 2003; Surmeier *et al*., 2017; Weinshenker, 2018), and noradrenergic treatments have been shown to modulate cognitive functions that are impaired in Parkinson’s disease, including response inhibition (Eagle *et al*., 2008; Vazey and Aston-Jones, 2012; Kehagia *et al*., 2014; Ye *et al*., 2015; Rae *et al*., 2016).

Response inhibition deficits are a well-documented feature of Parkinson’s disease, ranging from subclinical impairments to extreme impulsive behaviours (Gauggel *et al*., 2004, Obeso *et al*., 2011a; O’Callaghan *et al*., 2013; Nombela *et al*., 2014; Napier *et al*., 2015). Impulsivity is clear in the florid ‘impulse control disorders’ that are exacerbated by dopaminergic therapy (Weintraub *et al*., 2010). However, milder impulsivity is common in the absence of an impulse control disorder, including impairments in the ability to cancel an inappropriate action. Neurodegeneration of fronto-striatal circuits, including the subthalamic nucleus and its inputs, contributes to this impairment in Parkinson’s disease (O’Callaghan *et al*., 2013; Jahanshahi *et al*., 2014; Mosley *et al*., 2019), while pharmacological modulation of these circuits offers a tractable route to restorative treatment (Marsh *et al*., 2009; Kehagia *et al*., 2014; Ye *et al*., 2015; Rae *et al*., 2016).

The locus coeruleus-noradrenaline system modulates the stimulus detection and behavioural re-orienting required for rapid action cancellation (Robbins and Arnsten, 2009; Bari and Robbins, 2013). Phasic activation in the locus coeruleus – the brain’s main source of noradrenaline – occurs in response to salient events, and its activity is tightly time-locked to task-relevant responses (Aston-Jones and Cohen, 2005). Highly collateralised projections from the locus coeruleus enable release of noradrenaline in multiple brain regions, altering the gain, or responsivity, of target neurons. The action of noradrenaline at multiple targets can interrupt and reconfigure network architecture, promoting a change in goal-directed behaviour (Bouret and Sara, 2005; Zerbi *et al*., 2019). This locus coeruleus-noradrenaline function directly supports rapid action cancellation. In healthy adults and in preclinical models, pharmacologically increasing noradrenaline levels with the reuptake inhibitor atomoxetine improves action cancellation, as measured on stop-signal tasks (Chamberlain, 2006; Robinson *et al*., 2008; Chamberlain *et al*., 2009; Bari *et al*., 2011).

Atomoxetine selectively inhibits presynaptic noradrenaline transporters, resulting in a three-fold increase in extracellular levels of noradrenaline in the prefrontal cortex (Bymaster *et al*., 2002). It is currently licensed for treating behavioural and cognitive symptoms associated with attention deficit hyperactivity disorder. However, experimental psychopharmacological studies indicate it may be of value in some patients with Parkinson’s disease, by increasing activity and connectivity in the fronto-striatal ‘stopping network’ (Chamberlain *et al*., 2009; Bari *et al*., 2011; Rae *et al*., 2016). The stopping network includes the inferior frontal gyrus and presupplementary motor area, and their excitatory connection with the subthalamic nucleus which, via the globus pallidus, increases inhibition over thalamocortical output (Aron, 2011; Rae *et al*., 2014). Within this network, noradrenaline increases cortical excitability (McGinley *et al*., 2015), functional connectivity (Eldar *et al*., 2013) and network integration (Shine *et al*., 2018). The prefrontal regions also provide descending input to modulate the locus coeruleus (Arnsten and Goldman-Rakic, 1984; Jodo *et al*., 1998), by which prefrontal noradrenaline can influence locus coeruleus activity. Atomoxetine alters locus coeruleus firing patterns to increase the phasic-to-tonic ratio, making the locus coeruleus more responsive to task-relevant stimuli (Bari and Aston-Jones, 2013).

The potential for atomoxetine to modulate the locus coeruleus-noradrenaline function and improve response inhibition holds therapeutic promise in Parkinson’s disease. Previous work using stop-signal tasks in Parkinson’s disease demonstrated that atomoxetine can improve response inhibition and enhance its attendant stopping network activation (Kehagia *et al*., 2014; Ye *et al*., 2015, 2016; Rae *et al*., 2016). However, there was considerable heterogeneity in treatment response. To accurately predict the patients who would benefit from noradrenergic therapy remains a critical goal for atomoxetine treatment to be considered therapeutically and to design the necessary clinical trials with stratified patient selection (Matthews *et al*., 2014; Ye *et al*., 2016).

Here, we test the hypothesis that structural integrity of the noradrenergic locus coeruleus explains the improvements in response inhibition following atomoxetine. This is now possible through recent developments in ultra-high field 7T imaging of the locus coeruleus (Priovoulos *et al*., 2018; Betts *et al*., 2019; Ye *et al*., 2020). We tested this hypothesis by combining quantification of the locus coeruleus by 7T MRI with an acute psychopharmacological challenge, and measuring response inhibition using the stop-signal reaction time.

## Methods

### Participants

Nineteen people with idiopathic Parkinson’s disease were recruited via the University of Cambridge Parkinson’s disease research clinic and the Parkinson’s UK volunteer network. They met the United Kingdom Parkinson’s Disease Society Brain Bank criteria and were not demented based on MDS criteria for Parkinson’s disease dementia (Emre *et al*., 2007) nor on the mini-mental state examination (score >26) (Martinez-Martin *et al*., 2011). They were aged between 50-80 years, with Hoehn and Yahr stages 1.5-3, and had no contraindications to 7T MRI or atomoxetine. None had current impulse control disorders, based on clinical impression and the Questionnaire for Impulsive-Compulsive Disorders in Parkinson’s Disease (QUIP-Current Short) screening tool (Weintraub *et al*., 2009). Levodopa equivalent daily dose (LEDD) scores were calculated (Tomlinson *et al*., 2010). Twenty-six age-, sex- and education matched healthy controls were recruited from local volunteer panels. Control participants were screened for a history of neurological or psychiatric disorders, and no controls were using psychoactive medications. The study was approved by the local Ethics Committees and all participants provided written informed consent according to the Declaration of Helsinki. Demographic details and clinical characteristics are provided in Table 1 and the Supplementary Material.

**Table 1:** *Demographics and clinical assessments of participants in their normal on medication state. Data are presented as mean (SD). Comparisons of patient and control groups were performed with independent samples t-tests or contingency tables as appropriate*.

|  |  | PD | Controls | BF | *p* |
| --- | --- | --- | --- | --- | --- |
| Age (years) |  | 67.11 (7.05) | 65.35 (5.32) | 0.43 | .368 |
| Education (years) |  | 14.05 (2.27) | 14.65 (3.10) | 0.37 | .457 |
| Male / Female |  | 15 / 4 | 15 / 11 | 0.98 | .240 |
| MMSE |  | 29.47 (0.70) | 29.77 (0.51) | 0.87 | .128 |
| MoCA |  | 28.11 (1.76) | 28.58 (1.39) | 0.45 | .340 |
| ACE-R | Total Score | 94.89 (3.71) | 97.58 (3.16) | **4.16** | **.015** |
|  | Attention & Orientation | 17.84 (0.37) | 17.96 (0.20) | 0.64 | .216 |
|  | Memory | 23.68 (1.97) | 25.04 (1.18) | **7.00** | **.013** |
|  | Fluency | 12.00 (2.08) | 12.81 (1.60) | 0.71 | .167 |
|  | Language | 25.84 (0.50) | 25.88 (0.43) | 0.31 | .768 |
|  | Visuospatial | 15.63 (0.50) | 15.81 (0.63) | 0.45 | .302 |
| MDS-UPDRS | I: Nonmotor experiences | 9.00 (4.18) |  |  |  |
|  | II: Motor experiences | 12.63 (4.26) |  |  |  |
|  | III: Motor Examination | 28.42 (11.60) |  |  |  |
|  | IV: Motor Complications | 0.47 (0.96) |  |  |  |
|  | Total Score | 50.58 (17.20) |  |  |  |
| Hoehn and Yahr stage |  | 2.26 (0.45) |  |  |  |
| Disease duration (years) |  | 4.15 (1.72) |  |  |  |
| Levodopa equivalent daily dose (mg/day) |  | 644.55 (492.81) |  |  |  |

### Study procedure

Participants with Parkinson’s disease were tested across three sessions. Firstly, they underwent MRI scanning and clinical assessment, including the Movement Disorders Society Unified Parkinson’s Disease Rating Scale (MDS-UPDRS; (Goetz *et al*., 2008)), mini-mental state examination (MMSE), Montreal cognitive assessment (MoCA; (Nasreddine *et al*., 2005)) and the revised Addenbrooke’s cognitive examination (ACE-R; (Mioshi *et al*., 2006)).

On the second and third sessions, patients completed a double-blind randomised placebo-controlled crossover study, with 40 mg of oral atomoxetine or placebo. Drug/placebo order was randomly permuted in groups of six successive recruits. The visits were ≥ 6 days apart (mean 7.4 days; standard deviation 1.7 days; range 6-14 days). Blood samples were taken two hours after administration of drug/placebo, to coincide with predicted peak plasma concentration of atomoxetine after a single oral dose (Sauer *et al*., 2005). Mean plasma concentration (Teichert *et al*., 2020) was 261.32 ng/mL after atomoxetine (standard deviation 117.33 ng/mL, range 90.92-595.11 ng/mL) and 0 ng/mL after placebo. After the blood sample, patients commenced an experimental task battery that included a stop-signal response inhibition task. Supine/lying and upright blood pressure and pulse rate measures were monitored three times across the session (on arrival, two hours post tablet administration, on completion of testing). To monitor any changes in subjective feelings following the drug/placebo, prior to tablet administration and two hours post, we administered a set of 16 visual analogue scales (VAS) rating current mood and arousal levels. All sessions and MRI scanning were conducted with patients on their regular anti-parkinsonian medications and at a similar time of day.

Control participants were tested in one session to provide normative data on the task, in which they underwent MRI scanning and completed the same experimental task battery as the patients. The control group did not undergo the drug/placebo manipulation. Both the patient and control groups completed a set of self- and informant-rated questionnaires to assess mood and behaviour, which are reported in detail in the Supplementary Material.

### Stop-signal task

We used a stop-signal task to measure response inhibition. This paradigm involves a two-choice reaction time (RT) ‘go’ task that is occasionally interrupted by a ‘stop signal’, which requires the initiated response to be cancelled (Figure 1A). On go trials, a left- or right-pointing black arrow was presented on the screen, and participants indicated its orientation by pressing a left or right response button. On stop trials, the arrow changed colour from black to red at the same time as a tone (i.e., the stop-signal), after a short and variable delay (i.e., the stop-signal delay, SSD). Participants were instructed to inhibit any imminent response if the arrow became red. The length of the SSD was varied across stop trials using a staircase method to target a stop accuracy of 50%. The SSD ranged from 50 ms to 1500 ms and increased or decreased by 50 ms after a successful or failed stop trial, respectively. On no-go trials, the SSD was set to zero. The task consisted of 4 blocks of 140 trials each, including 110 go trials, 10 no-go trials (very low commission error rate), and 20 stop trials per block (approximately 50% commission error rate). The first 20 trials of each block were go trials, to compute a starting value for the SSD (mean RT minus 200 ms). The remaining trials within each block were pseudorandomly interleaved, with the constraints that there could be no more than seven consecutive go trials, and no more than two consecutive no-go or stop trials. At the start of each trial, a fixation cross was presented for 500 ms.

**Figure 1.**
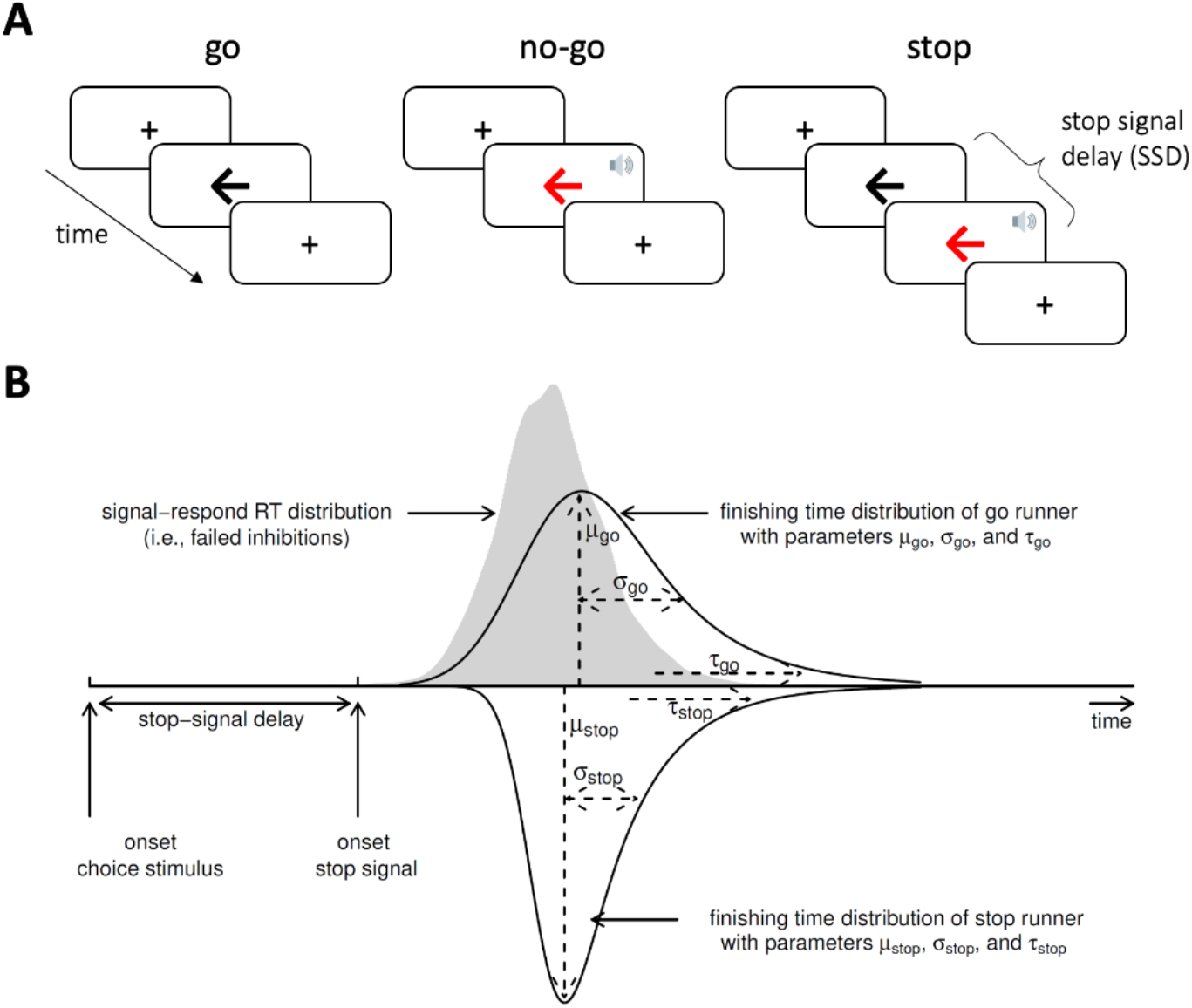
*Design of the stop signal task (A) and ex-Gaussian race model of response inhibition (B). (A) In the stop-signal go/no-go task, participants respond as quickly and accurately as possible to the direction of a black arrow (go trials). Occasionally, this task is interrupted by a stop signal (red arrow and beep tone), which requires any imminent response to be inhibited. For no-go trials, the stop signal is presented immediately after the fixation cross. For stop trials, the stop signal is presented after an initial go stimulus, with a short and variable delay. (B) The ex-Gaussian race model characterises task performance as a race between three competing processes or ‘runners ’: One stop process, and two go processes that match or mismatch the go stimulus. The finishing times of each process are assumed to follow an ex-Gaussian distribution. Successful inhibition in no-go or stop trials occurs when the stop process finishes before both go processes. A correct go response occurs when the matching go process finishes before the mismatching go process and stop process. For simplicity, the finishing time distribution of the mismatching go process is not illustrated. Acknowledgements: The speaker symbol in Figure 1A was copied from the Twitter emoji library, available at https://twemoji.twitter.com/under a CC-BY 4.0 license (https://creativecommons.org/licenses/by/4.0/). Figure 1B was copied from Heathcote *et al*. (2019), available at https://flic.kr/p/24g3sip under a CC-BY 2.0 license (https://creativecommons.org/licenses/by/2.0/)*.

Participants were given standardised instructions at the start of the experiment. They were asked to respond as ‘quickly and accurately as possible’, and were discouraged from strategically slowing down in anticipation of a stop signal (Verbruggen *et al*., 2019). After these instructions, they were given a practice block of 25 trials (20 go trials, 2 no-go trials, and 3 stop trials). The experimenters verified that the participant understood the task, and if necessary the practice block was repeated. The practice data was not analysed further.

### Ex-Gaussian race model of response inhibition

We used a Bayesian parametric model of the stop-signal task to infer the latency of the unobservable stop response – the stop-signal reaction time (SSRT; (Matzke *et al*., 2013, 2019). This model assumes a race between three independent processes: one corresponding to the stop process, and two corresponding to go processes that match or mismatch the go stimulus (Figure 1B). For a given stop trial, successful inhibition occurs when the stop process finishes before both go processes. For a given go trial, a correct response occurs when the matching go process finishes before the mismatching go process. The finish time distribution of the stop process is inferred by estimating the RT distribution of unsuccessful stop trials (i.e., signal respond RTs). Specifically, the signal respond RT distribution is assumed to be a right-censored go RT distribution, where the censoring point for a given stop trial is drawn from the finish time distribution of the stop process (see (Matzke *et al*., 2013) for details).

The model assumes that the finish times of the stop and go processes follow an ex-Gaussian distribution, which is a positively skewed unimodal distribution that is commonly used to describe RT data (Ratcliff, 1979; Heathcote *et al*., 1991). Thus, for each process, we estimated the three parameters of the ex-Gaussian distribution: The mean μ and standard deviation σ of the Gaussian component, and the mean (i.e., inverse rate) τ of the exponential component.

We additionally estimated two parameters that represent the probability that the stop and go processes failed to start, referred to as “trigger failure” and “go failure”, respectively (Matzke *et al*., 2019). These attentional failures are common in both healthy participants (Matzke *et al*., 2017b; Skippen *et al*., 2019) and in clinical cohorts (Matzke *et al*., 2017a; Weigard *et al*., 2019), and if not modelled can severely bias estimation of the stop process (Band *et al*., 2003; Matzke *et al*., 2019; Skippen *et al*., 2019). Prior to fitting the model, we removed implausibly short (< 0.25 s) or long (> 4.5 s) RTs, as well as go RTs more extreme than ±2.5 standard deviations from the participant’s mean (Matzke *et al*., 2013).

We used Markov Chain Monte Carlo (MCMC) sampling to estimate the posterior distributions of the parameters. The parameters were estimated hierarchically, such that parameters for a given participant were sampled from corresponding group-level distributions. We fitted this hierarchical model separately for the control group, the Parkinson’s disease group on placebo, and the Parkinson’s disease group on atomoxetine. We placed the same set of prior distributions on the group-level parameters for each of these three groups. The prior distributions were identical to those suggested by the model developers (Heathcote *et al*., 2019), except for slightly higher prior mean values for μ_go-match_ (1.5 s), μ_go-mismatch_ (1.5 s) and μ_stop_ (1 s), to account for slower RT in older age (see Supplemental Material for full list of priors). The model MCMC sampling initially ran with 33 chains (i.e., three times the number of parameters), with thinning of every 10th sample and a 5% probability of migration. Model convergence was assessed with the potential scale reduction statistic *Ṙ* (<1.1 for all parameters), and with visual inspection of the time-series plots of the chains. After this, an additional 500 iterations for each chain were run to create a final posterior distribution for each parameter. To assess the model’s goodness of fit, the observed data was compared to simulated data generated from the model’s posterior predictive distribution (see Supplementary Material Figures 4 – 6).

The primary outcome of interest, SSRT, was computed as the mean of the ex-Gaussian finish time distribution of the stop process, which is given by μ_stop_ + τ_stop_. We repeated this for each MCMC sample to approximate a posterior distribution of SSRT. This approach was also used to approximate a posterior distribution of go RT (μ_go-match_ + τ_go-match_).

### Statistical analysis

The go error rate and stop accuracy rate served as basic descriptive statistics for stop-signal task performance. We defined the go error rate as the proportion of go trials with an incorrect response, including commission errors (responses that mismatch the arrow direction) and omission errors (missing responses). The stop accuracy rate was defined as the proportion of stop trials with successfully inhibited responses (missing responses). For both outcomes, we examined differences between groups (Parkinson’s disease placebo vs. controls) and drug conditions (Parkinson’s disease placebo vs. atomoxetine) using independent and paired samples *t*-tests respectively.

We then examined the group- and participant-level parameter estimates from the ex-Gaussian race model described above. For group-level inference, we examined the posterior distributions of the group-level means of SSRT and go RT. For a given posterior distribution, we took the median as the posterior estimate, and the 95% quantile interval (QI) as the range of most credible values. We also obtained posterior distributions for contrasts of interest (Parkinson’s disease placebo vs. controls; Parkinson’s disease placebo vs. atomoxetine) by subtracting the sets of MCMC samples of the two groups under consideration. That is, for a given parameter, we computed the difference between the two groups for each MCMC sample, thereby yielding an approximate posterior distribution of the difference (Kruschke, 2013).

To test for individual differences in the effect of atomoxetine on SSRT and go RT, we extracted the medians of the participant-level posterior distributions of SSRT and go RT from the placebo and atomoxetine model fits. We hypothesised that the effect of atomoxetine would depend on the integrity of the locus coeruleus, indexed by the contrast-to-noise ratio (CNR; described below). Therefore, the posterior parameter estimates were entered as the dependent variable with drug condition (placebo vs. atomoxetine), CNR, and their interaction as fixed effects, allowing the intercept to vary across participants (random effect). We additionally included a fixed effect of session (first vs. second) as a covariate of no interest. Taking the analysis of SSRT as an example, the model was specified in R formula syntax as follows: `SSRT ~ drug * CNR + session + (1 | subject)`.

For linear models, we report both frequentist and Bayes factor (BF) analyses for hypothesis testing, with a significance threshold of p =.05 (two-sided) for frequentist analyses. We present the BF for the alternative hypothesis over the null hypothesis (i.e., BF_10_), such that BF > 1 indicates relative evidence for the alternative hypothesis, and BF > 3 indicates “positive evidence” for the alternative hypothesis (Kass and Raftery, 1995). All BF analyses used the default ‘JZS’ prior on the effect size under the alternative hypothesis (Rouder *et al*., 2009, 2012). To test for specific fixed effects in linear mixed models, we obtained p-values using the Kenward-Roger method, and BFs through Bayesian model averaging by estimating the change from prior to posterior inclusion odds (inclusion BF). In other words, this model-averaged BF indicates how much more likely the data are under model variants that include a given fixed effect, compared to model variants that exclude the fixed effect (Hinne *et al*., 2020).

Due to technical issues, stop-signal task data was missing for one patient’s placebo session and for another patient’s atomoxetine session. We nevertheless included these two participants in the linear mixed model analyses of the within-subject effect of atomoxetine, as participants were treated as a random effect. However, excluding these two participants did not meaningfully change any of the following results.

### Software and Equipment

The stop-signal task was implemented in MATLAB R2018b using the Psychophysics Toolbox extensions (Version 3; (Kleiner *et al*., 2007)). Participants responded using a two-button response box. The ex-Gaussian model fitting was performed with the Dynamic Models of Choice toolbox (Heathcote *et al*., 2019), implemented in R (version 3.6.1, R Core Team, 2019). Further statistical analyses in R used the ‘tidyverse’ (Wickham *et al*., 2019) and ‘tidybayes’ (Kay, 2020) packages for data organisation and visualisation, the ‘afex’ package (Singmann *et al*., 2020) for ANOVA and linear mixed model fitting with the ‘emmeans’ package (Lenth *et al*., 2020) used for post hoc comparisons, and the ‘BayesFactor’ (Morey *et al*., 2018) and ‘bayestestR’ (Makowski *et al*., 2019) packages for Bayes Factor analysis.

### MRI acquisition

All patients and controls underwent MR imaging. Two controls were excluded from further imaging analysis due to incidental structural abnormalities. MR images were acquired with a 7T Magnetom Terra scanner (Siemens, Erlangen, Germany), using a 32-channel receive and circularly polarised single-channel transmit head coil (Nova Medical, Wilmington, USA). We used a 3-D high-resolution magnetisation transfer-weighted turbo flash (MT-TFL) sequence for imaging the locus coeruleus (based on (Priovoulos *et al*., 2018)). 112 axial slices were used to cover both the midbrain and the pontine regions. The sequence applied a train of 20 Gaussian-shape RF-pulses at 6.72 ppm off resonance, 420° flip-angle, followed by a turbo-flash readout (TE = 4.08 ms, TR = 1251 ms, flip-angle = 8°, voxel size = 0.4 x 0.4 x 0.5 mm^3^, 6/8 phase and slice partial Fourier, bandwidth = 140 Hz/px, no acceleration, 14.3%- oversampling, TA ~ 7 min). For each subject, the transmit voltage was adjusted based on the average flip angle in the central area of the pons obtained from a B1 pre-calibration scan. The MT-TFL sequence was repeated twice and averaged offline to improve signal-to-noise ratio. An additional scan (MT-off) was acquired with the same parameters as above but without the off-resonance pulses. A high resolution isotropic T1-weighted (T1-w) MP2RAGE image was also acquired sagittally for anatomical coregistration using the UK7T Network harmonised protocol (Clarke *et al*., 2020): TE = 2.58 ms, TR = 3500 ms, BW = 300 Hz/px, voxel size = 0.7 x 0.7 x 0.7 mm^3^, FoV = 224 x 224 x 157 mm^3^, acceleration factor (A>>P) = 3, flip angles = 5/2° and inversion times (TI) = 725/2150 ms for the first/second images.

### Image processing and coregistration pipeline

Image processing and coregistration was based on the pipeline described in (Ye *et al*., 2020). The Advanced Normalization Tools (ANTs v2.2.0) software and in-house Matlab scripts were used for image pre-processing and the standardisation of MT images. MT images were first N4 bias field corrected for spatial inhomogeneity (number of iterations at each resolution level: 50×50×30×20, convergence threshold: 1×10^-6^, isotropic sizing for b-spline fitting: 200) (Tustison *et al*., 2010) then averaged using the customised antsMultivariateTemplateConstruction2 function for improvements in signal-to-noise ratio. The T1-w MP2RAGE data were generated offline from the complex images (Clarke *et al*., 2020). T1-w skull-stripped images were obtained after tissue type segmentation and reconstruction using SPM12 (v7219) (http://www.fil.ion.ucl.ac.uk/ spm/software/ spm12/).

The pre-processed MT-weighted and T1-w images were then entered into a T1-driven, cross modality coregistration pipeline to warp the individual MT and MT-off images to the isotropic 0.5 mm ICBM152 (International Consortium for Brain Mapping) T1-w asymmetric template (Fonov *et al*., 2011). The individual T1-w images were first coregistered to the MT image with rigid only transformation. The MT-off image was used as the intermediate step for bridging the two modalities because the MT-off image shares similar tissue-specific contrasts with both T1-w MP2RAGE and MT-on images.

In parallel, an unbiased study-wise T1-w structural template was created using individual skull-stripped T1-w images from all controls and patients. Native T1-w images were rigid and affine transformed, followed by a hierarchical nonlinear diffeomorphic step at five levels of resolution, repeated by six runs to improve convergence. Max iterations for each resolution from the coarsest level to the full resolution were 100×100×70×50×20 (shrink factors: 10×6×4×2×1, smoothing factors: 5×3×2×1×0 voxels, gradient step size: 0.1 mm). The Greedy Symmetric Normalisation (SyN) was adopted for the transformation model of the deformation step (Avants *et al*., 2008). The resulting T1-w group template was then registered to the standard ICBM152 T1-w brain following the similar rigid-affine-SyN steps at four resolution levels (max iterations: 100×70×50×50, convergence threshold: 1×10^-6^, shrink factors: 8×4×2×1, smoothing factors: 3×2×1×0 voxels). For all the above registration steps, cross-correlation was used for similarity metrics estimation as it performs better for linear and non-linear components during intra-modality registration. Four steps of deformations were estimated as follows (in order): MT-off to MT, T1-w to MT-off, T1-w to T1-w group template and T1-w group template to ICBM152 T1-w template. These parameters were then used as the roadmap for MT image standardisation to the ICBM brain in one step. A trilinear interpolation method was selected to preserve the absolute location and relative contrast of the signal.

### Independent probabilistic locus coeruleus atlas creation

To facilitate accurate extraction of the locus coeruleus signal we created a study-specific unbiased locus coeruleus atlas (See Figure 2A). To do this, we used an independent sample of 29 age- and education matched healthy controls (13 female; age mean (SD) = 67 (8.2), age range = 52-84) collected under the same neuroimaging protocol. We used a validated pipeline for locus coeruleus atlas construction described in (Ye *et al*., 2020). Briefly, for each axial slice on the rostrocaudal extent, the locations of the left and right locus coeruleus were determined using a semi-automated segmentation method. A threshold was defined as five standard deviations above the mean intensity in the central pontine reference region. After applying the threshold, locus coeruleus voxels on axial planes were automatically segmented into binarised images then averaged to construct a probabilistic atlas. The independent locus coeruleus atlas generated for this study had very high similarity in the spatial distribution of probabilities and contours relative to the validated 7T locus coeruleus atlas in (Ye *et al*., 2020) (See Supplementary Figure 1).

**Figure 2.**
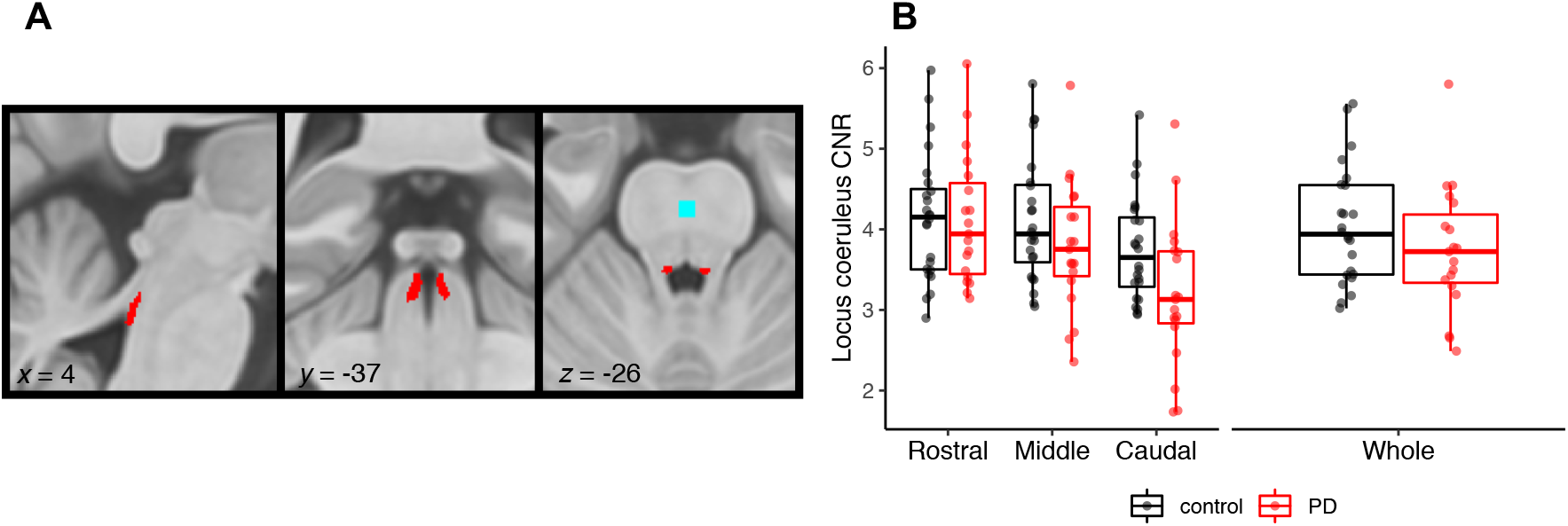
*A) Study specific locus coeruleus atlas, also showing the reference region (blue) in the central pons; b) Contrast-to-noise ratio (CNR) for the locus coeruleus subdivisions and whole structure in Parkinson’s disease patients vs. controls (note, left and right locus coeruleus are combined)*.

### Locus coeruleus signal extraction

As a measure of locus coeruleus integrity, we quantified contrast by establishing the CNR with respect to a reference region in the central pons (see Figure 2A). A CNR map was computed voxel-by-voxel on the average MT image for each subject using the signal difference between a given voxel (V) and the mean intensity in the reference region (MeanREF) divided by the standard deviation (SD_REF_) of the reference signals 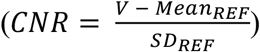. CNR values were extracted bilaterally on the CNR map by applying the independent locus coeruleus probabilistic atlas (5% probability version). We computed mean CNR values for the rostral, middle and caudal portions of the left and right locus coeruleus. As an index of locus coeruleus integrity to incorporate with the stop-signal task analysis, we combined the left and right locus coeruleus and averaged across the whole structure.

### Data availability

Code and data to reproduce manuscript figures, statistical analyses and stop-signal task modelling is freely available through the Open Science Framework [OSF link to be added].

## Results

### Behavioural results

As shown in Table 1, the patient and control groups were matched in terms of age, years of education, sex ratio, MMSE and MoCA (all p values >.050; all BFs < 1). Patients had a significantly lower ACE-R total score (p =.015, BF = 4.16), and lower memory subscale (p =.013, BF = 7.00). There were no group differences on any of the self-reported questionnaires assessing impulsivity, anxiety, depression and behavioural measures, with the exception of the Motivation and Energy Inventory where the patient group had significantly lower scores for the physical subscale compared to controls (p =.010, BF = 6.18; Supplementary Material Table 1).

Within the drug and placebo sessions, there was some evidence of increased pulse rates and raised blood pressure on atomoxetine, although this was not considered clinically relevant and was not consistently observed across all supine and upright measures. Importantly, there was no change in subjective ratings of mood and arousal levels within the sessions. These analyses are described in detail in the Supplementary Material.

### Locus coeruleus integrity

Figure 2B shows comparisons of locus coeruleus CNR between the patients and controls. When comparing across the whole structure, the groups were not significantly different (*t*_(36.98)_ = 1.27, *p* = 0.21, BF = 0.58). Comparing across the rostral, middle and caudal subdivisions, there wasa main effect of subdivision *(F*_(1.27_, _52.22)_ = 54.57, *p* <.001; BF = 5.28 x 10^10^). This was driven by CNR in the caudal portion being significantly lower than both the middle (*t*_(82)_ = 7.37, *p* <.0001) and rostral (*t*_(82)_ = 10.10, *p* <.0001) portions. There was a significant group by subdivision interaction (*F*_(1.27, 52.22_) = 7.89*,p =* 004; BF = 32.98). This reflected significantly lower CNR values in the caudal portion for patients relative to controls (*t*_(50.1)_ = 2.23*,p =* 0.026), whereas the groups did not differ for the middle (*t*_(50.1)_ = 1.32, *p* = 0.193) or rostral (*t*<u>_(50.1)_</u> = 0.140 *,p =* 0.889) portions of the locus coeruleus.

### Stop-signal task performance

In keeping with the tracking algorithm, the stop accuracy for the Parkinson’s disease group on placebo (Figure 3 A; M = 0.48, SD = 0.15) was not significantly different from controls (M = 0.58, SD = 0.18; *t*_(40.79)_ = 1.90, *p* =.065, BF = 0.35). Across the Parkinson’s disease patients, group-wise stop accuracy on atomoxetine (M = 0.45, SD = 0.13) was not significantly different from the placebo session *(t*_(16)_ = 0.88, *p* =.39, BF = 0.35). The go error rate approached zero for most participants, yielding a skewed distribution bounded at zero (Figure 3D), and was therefore logit transformed prior to analysis (Warton and Hui, 2011). The logit go error rate was slightly higher in the Parkinson’s disease group on placebo (M = -4.29, SD = 1.23) than in controls (M = -5.04, SD = 0.93; *t*_(29.94)_ = -2.19, *p* =.037, BF = 0.46). Within the Parkinson’s disease group, the logit go error rate was marginally reduced on atomoxetine (M = -4.75, SD = 1.22) compared to placebo (*t*_(16)_ = 2.23, *p* =.041, BF = 1.73). However, we note that the BF for both these tests fell below conventional thresholds for positive evidence (i.e., BF > 3), and these effects on go error rate should therefore be regarded as ‘anecdotal’ at the group level.

**Figure 3.**
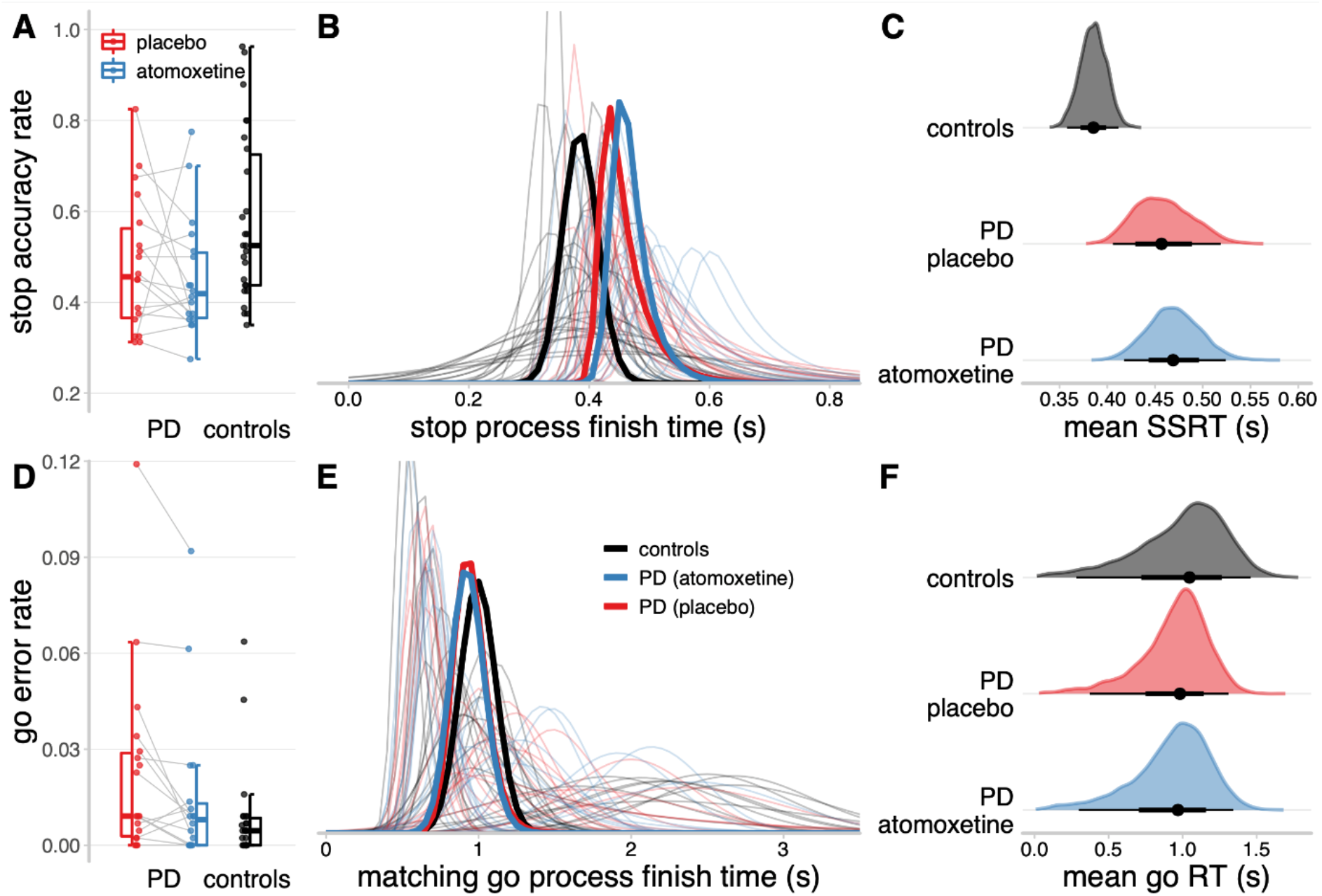
*Descriptive statistics and ex-Gaussian model estimates of stop-signal task performance. A, D: Proportions of successful stop trials (A) and incorrect go responses (D). B, E: Ex-Gaussian finish time distributions of the stop process (B) and matching go process (E). Bold lines represent group-level mean distributions; thin lines represent individual participants. The mean of a given ex-Gaussian distribution was taken as the SSRT (stop process) or go RT (matching go process). C, F: Posterior distributions of group-level mean SSRT (C) and go RT (F). The black dots represent the medians; the thick black line segments represent the 66% quantile intervals; and the thin black line segments represent the 95% quantile intervals*.

### Ex-Gaussian model estimates of SSRT

The hierarchical Bayesian estimates of the ex-Gaussian finish time distributions for the stop and matching go processes are shown in Figures 3B and 3E respectively. The stop process finish times tended to be faster for the control group than the Parkinson’s disease group. Indeed, the posterior distribution of group-level mean SSRT (Figure 3C) was lower for the control group (median = 0.39 s, 95% QI: [0.36, 0.41]) than the Parkinson’s disease group on placebo (median = 0.46 s, 95% QI: [0.41, 0.52]), and this group difference in SSRT was reliably different from zero (Δgroup median = 0.07 s, 95% QI: [0.01, 0.14]). The mean SSRT for the Parkinson’s disease group on atomoxetine (median = 0.47 s, 95% QI: [0.42, 0.52]) was comparable to the placebo session (Δdrug median = 0.01 s, 95% QI: [-0.07, 0.09]).

For the matching go process, the distributions of finish times varied widely across participants, but the group-level distributions were highly similar. The posterior distributions of group-level mean go RT (Figure 3F) did not differ between the control group (median = 1.05 s, 95% QI: [0.28, 1.46]), Parkinson’s disease group on placebo (median = 0.98 s, 95% QI: [0.37, 1.31]; Δgroup median = -0.07 s, 95% QI: [-0.78, 0.77]), and Parkinson’s disease group on atomoxetine (median = 0.97 s, 95% QI: [0.30, 1.34]; Δdrug median = -0.01 s, 95% QI: [-0.72, 0.67]). There were also no mean differences between groups or drug conditions for the attentional failure parameters, trigger failure and go failure (Supplementary Figure 7).

### Locus coeruleus integrity and atomoxetine-induced changes in SSRT

Although there was no group-wise effect of atomoxetine on the Parkinson’s disease group in terms of their mean SSRT, we predicted that the effect of atomoxetine would depend on individual differences in locus coeruleus integrity, as indexed by the CNR. We confirmed a significant interaction effect between the drug condition and locus coeruleus CNR on the participant-level estimates of SSRT (Figure 4A; β = 0.27, *F*_(1, 14.61)_ = 14.61, *p* =.002; BF = 11.56). This interaction effect did not meaningfully change when including clinical covariates such as age, disease severity, atomoxetine plasma level and dopaminergic medication, as both frequentist and Bayesian model selection procedures indicated that such covariates did not significantly improve the model fit (for details see Supplementary Material). There was also a main effect of session (β = 0.25, *F*_(1, 14.53)_ = 13.33, *p* =.002; BF = 2.58), reflecting slightly shorter SSRTs for the second session compared to the first, regardless of the drug condition. There were no significant main effects of drug condition (β = -0.05, *F*_(1, 14.21)_ = 0.70, *p* =.416; BF = 0.37) or locus coeruleus CNR (β = 0.11, *F*_(1, 14.61)_ = 0.266, *p* =.613; BF = 0.55) on SSRT.

**Figure 4.**
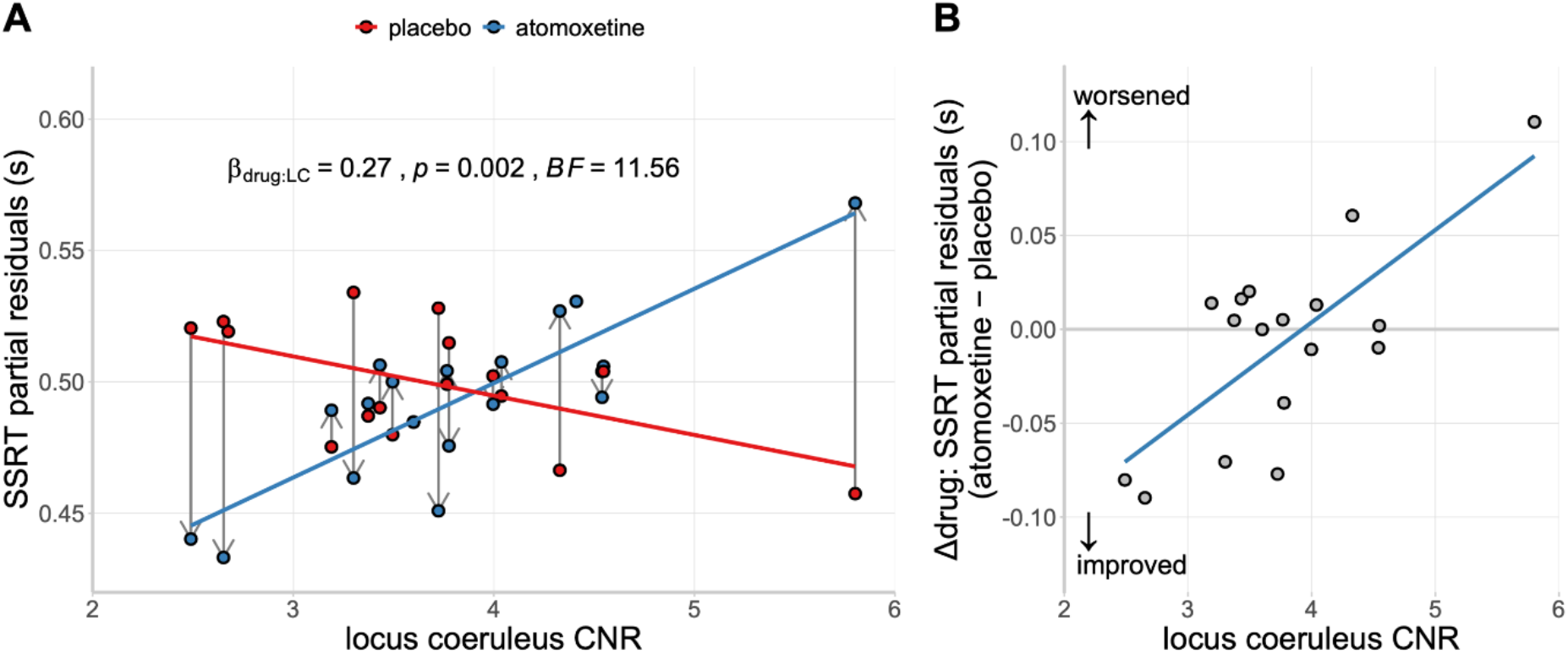
*(A) SSRT estimates as a function of drug condition and locus coeruleus contrast-to-noise ratio (CNR). Within-subject change in SSRT from placebo to atomoxetine is illustrated with vertical grey arrows. (B) Relationship between locus coeruleus CNR and the drug-induced change in SSRT. Note: For visualisation purposes, the SSRT estimates were adjusted for the fixed effect of session and random effect of participants (i.e., partial residuals)*.

To further understand the drug × locus coeruleus CNR interaction, we examined the relationship between locus coeruleus CNR and the drug-induced change in SSRT (Δdrug: atomoxetine – placebo), adjusted for the main effect of session. This relationship was strongly positive, suggesting that patients with lower locus coeruleus CNR have a greater reduction in SSRT after atomoxetine (Figure 4B; *r*_(15)_ = 0.73, *p* <.001; BF = 32.70).

There was no interaction effect between drug condition and locus coeruleus CNR on the participant-level estimates of go RT (β = 0.01, *F*_(1, 16.37)_ = 0.00, *p* =.971; BF = 0.47), stop accuracy rate (β = -0.18, *F*_(1, 16.52)_ = 1.15, *p* =.300; BF = 0.58), or logit go error rate (β = -0.02, *F*_(1, 15.14)_ = 0.03, *p* =.865; BF = 0.46).

## Discussion

We show that improvements in response inhibition after atomoxetine are dependent on locus coeruleus integrity in people with Parkinson’s disease. Following a single dose of atomoxetine 40 mg, individuals with lower locus coeruleus integrity had a greater improvement in response inhibition (i.e., reduction in their stop-signal reaction time; SSRT). This result highlights the link between integrity of the noradrenergic locus coeruleus and action cancellation, which has previously been inferred from preclinical work and pharmacological manipulations. The finding also demonstrates the importance of baseline noradrenergic capacity in determining the response to atomoxetine, confirming the need to stratify patients for noradrenergic therapy. Locus coeruleus neuromelanin imaging would be a safe and affordable means to achieve this stratification.

Previous work in Parkinson’s disease showed that atomoxetine led to greater improvements in SSRT and enhanced activation in the stopping network in patients with more severe disease (Ye *et al*., 2015; Rae *et al*., 2016). Extending this work, we confirm that improved SSRTs under atomoxetine occurred in those patients with more severe locus coeruleus degeneration. This was in the context of a general impairment in response inhibition, as patients on placebo had longer SSRTs compared to controls. Our result suggests that a single 40 mg atomoxetine dose confers most benefit on individuals with a severe loss of noradrenergic capacity. In this way, noradrenergic replacement in patients with a compromised system may achieve restoration closer to normal levels and improve behaviour. Conversely, in patients with a less affected system the same dose may offer no appreciable benefit or even ‘overdose’ the system, leading to worse behaviour. This relationship is captured by the inverted-U shaped curve (known as a Yerkes-Dodson function) that is common across monoaminergic and cholinergic systems, whereby intermediate levels of neuromodulatory influence are associated with optimal performance, with too much or too little having deleterious effects on behaviour (Robbins, 2000; Aston-Jones and Cohen, 2005).

Baseline dependency in dose-response curves is well documented in relation to dopaminergic therapy (Rowe *et al*., 2008; Cools and D’Esposito, 2011), such that the optimal level of dopamine enhancement needed to improve behaviour depends on pre-existing dopamine levels. This has had critical implications for Parkinson’s disease therapy. Dopamine dosages titrated to restore levels in the severely depleted dorsal striatum and motor system circuitry effectively overdose the less affected ventral tegmental area, ventral striatum and associated limbic pathways (Cools *et al*., 2001), impairing aspects of learning and cognitive flexibility (Cools *et al*., 2007; MacDonald *et al*., 2011; Aarts *et al*., 2014). Our result suggests a similar baseline dependency for noradrenergic therapy, where optimal dosages needed for atomoxetine therapy may depend on the extent of degeneration in the locus coeruleus. This has important implications for optimising noradrenergic therapy in Parkinson’s disease, as patients could be stratified based on locus coeruleus integrity to inform appropriate dosages in clinical trials or personalised treatment (Ye *et al*., 2016).

Neuromelanin-sensitive magnetisation transfer imaging of the locus coeruleus represents a promising avenue to achieve this stratification. Although we have previously shown a relationship between disease severity (as measured by the UPDRS-III) and atomoxetine responsivity, such measures of motor function or disease duration may not be the most accurate measure of noradrenergic capacity. Whilst progressive degeneration of the locus coeruleus is expected over the disease course, this will vary widely across individuals and will reflect the disease phenotype. Neuropathological studies and neuromelanin imaging have shown more pronounced locus coeruleus degeneration in certain phenotypes, including those with cognitive impairment or dementia (Cash *et al*., 1987; Zweig *et al*., 1993; Li *et al*., 2019), depression (Wang *et al*., 2018), an akinetic-rigid syndrome (Paulus and Jellinger, 1991) and REM sleep behaviour disorder (García-Lorenzo *et al*., 2013; Sommerauer *et al*., 2017), relative to patients at equivalent disease stages. In our results we note that including the MDS-UPDRS-III as an index of disease severity did not meaningfully improve the model fit for the interaction between the drug condition and locus coeruleus CNR. This highlights the added value of locus coeruleus imaging, above and beyond disease severity metrics, to explain variations in atomoxetine responsivity.

Our locus coeruleus imaging identified that the greatest difference between controls and patients was in the caudal portion of the locus coeruleus. This has not previously been identified in Parkinson’s disease locus coeruleus imaging with 3T MRI, which limited analysis to the whole structure. While some neuropathology studies have noted comparable cell loss across the rostral-caudal extent of the locus coeruleus (Chan-Palay and Asan, 1989; German *et al*., 1992), others have reported more severe degenerative changes in the caudal segment (Bertrand *et al*., 1997).

Atomoxetine is known to increase extracellular noradrenaline levels across the brain via its actions at the noradrenaline transporter, in particular increasing levels in the prefrontal cortex by three-fold (Bymaster *et al*., 2002; Swanson *et al*., 2006). In the prefrontal cortex, atomoxetine may also increase extracellular dopamine levels (Bymaster *et al*., 2002). Due to the relative sparsity of dopamine transporters in the prefrontal cortex (Sesack *et al*., 1998), a portion of dopamine uptake is mediated by the noradrenaline transporter (Wayment *et al*., 2001; Seamans and Yang, 2004). Therefore, improved response inhibition under atomoxetine in Parkinson’s disease might potentially also reflect elevated prefrontal dopamine levels. Nevertheless, work in both rodents and humans implicates a selective link between noradrenergic transmission and action cancellation: increasing dopamine selectively does not affect the SSRT (Overtoom *et al*., 2003; Bari *et al*., 2009, Obeso *et al*., 2011b). The association we have shown between locus coeruleus integrity and the change in SSRT under atomoxetine suggests a direct link between the noradrenergic system and action cancellation in Parkinson’s disease.

Our results do not speak directly to atomoxetine’s mechanism of action in Parkinson’s disease. However, convergent evidence indicates that atomoxetine may increase efficiency in brain networks mediating response inhibition, via actions at the prefrontal cortex and the locus coeruleus (Bymaster *et al*., 2002; Bari and Aston-Jones, 2013). In Parkinson’s disease patients, atomoxetine has been shown to increase activity within and between regions of the stopping network, including the pre-supplementary motor area and inferior frontal gyrus (Ye *et al*., 2015; Rae *et al*., 2016). Locus coeruleus degeneration in Parkinson’s disease is accompanied by reduced noradrenaline levels in forebrain regions (Scatton *et al*., 1983; Cash *et al*., 1987; Pifl *et al*., 2012). As noradrenaline release facilitates reconfigurations of large-scale networks (Bouret and Sara, 2005; Zerbi *et al*., 2019), depletion of forebrain noradrenaline is likely to impact the rapid engagement of brain network activity that is necessary for successful action cancellation (Tsvetanov *et al*., 2018). In patients with greater locus coeruleus degeneration, which may be accompanied by decreased or dysfunctional modulation of prefrontal noradrenergic targets, efficiency of the stopping network may be reduced. Consequently, these patients show the greatest benefit from a drug that can increase levels of prefrontal noradrenaline and upregulate locus coeruleus function.

Given the role of noradrenaline in cognition and behaviour (Sara, 2009), noradrenergic dysfunction contributes to cognitive deficits beyond action cancellation in Parkinson’s disease. Optimising noradrenergic therapy therefore has potential to provide relief across a variety of non-motor symptoms (Oertel *et al*., 2019). Our results confirm the potential for stratified noradrenergic therapy in Parkinson’s disease, whereby the efficacy of these drugs varies across individuals depending on their baseline noradrenergic state. Locus coeruleus neuromelanin imaging may offer a marker of noradrenergic capacity that can be used to stratify patients to optimise successful outcomes in trials of noradrenergic therapy, and ultimately inform a more personalised treatment approach.

## Acknowledgments

We thank all volunteers and their families for their participation, all staff at the Wolfson Brain Imaging Centre and NIHR Cambridge Clinical Research Facility for their help with data collection, and members of the Cambridge Centre for Frontotemporal Dementia and Related Disorders for valuable suggestions and discussions.

